# GLP1R Variants and Polygenic Risk Underlie Heterogeneous Response to GLP-1 Receptor Agonists in Type 2 Diabetes

**DOI:** 10.64898/2026.07.29.26359248

**Authors:** Munichandra Babu Tirumalasetty, Vivian Hsing-Chun Wang, Mohammad Sarif Mohiuddin, Mayank Choubey, Rashu Barua, Donglan Stacy Zhang, Qing Robert Miao

## Abstract

**Objective:** To identify clinical and genetic factors associated with variation in glycemic response to glucagon-like peptide-1 receptor agonist (GLP-1RA) therapy among adults with type 2 diabetes, with a focus on common GLP1R variations, polygenic risk load, and pancreas-specific regulation annotation.

**Research Design and Methods:** We conducted a retrospective cohort study using electronic health record (EHR)-linked biobank data from the All of Us research workbench platform that included 5784 adults with type 2 diabetes who initiated GLP-1RA therapy. Baseline HbA1c was measured within 3 months before medication initiation, and follow-up HbA1c was measured after 3 months. The patients with type 2 diabetes were classified as good responders (HbA1c reduction ≥ 2.5 percentage point) or poor responders (HbA1c reduction <0.5 percentage point). Models adjusted for demographic characteristics, anthropometric and metabolic measures, blood pressure, body mass index (BMI), lipid profile, liver function tests, polygenic risk score, and GLP1R variant carrier status were compared between the two groups. Common GLP1R variations were further investigated for carrier frequency and associated HbA1c levels before and after medication use.

**Results:** The cohort included 3194 good responders and 2590 poor responders. Good responders were younger than poor responders (55.2 vs. 58.6 years) and had significantly higher glycemic improvement. HbA1c levels fell from 9.2% to 6.3% in good responders and 8.4% to 8.1% in poor responders, resulting in an absolute HbA1c reduction of 2.9% and 0.3%, respectively. Good responders also showed larger decreases in fasting glucose, BMI, systolic and diastolic blood pressure, triglycerides, total cholesterol, LDL cholesterol, and liver enzymes, as well as minor improvements in HDL-C. After multivariable adjustment, Poor responders had a greater T2D polygenic risk score (0.38 vs. 0.21), more GLP1R coding variant carrier status (10.1% vs. 8.0%), and a higher overall GLP1R variant burden (22.8% vs. 19.2%). Variant-level studies revealed rs2268650 and rs2003132 enrichment among poor responders, with negative post-treatment HbA1c patterns in carriers, whereas good-response carriers showed significant HbA1c improvement.

**Conclusions:** Response to GLP-1RA in T2D is associated with baseline clinical and metabolic status, as well as inherited genetic susceptibility, which includes common GLP1R variation and a larger polygenic risk burden. Integrating clinical and pharmacogenomic profiling may improve patient classification and provide insight into treatment failure in poor responders.

## Introduction

Type 2 diabetes (T2D) is a chronic and diverse metabolic illness that continues to cause significant morbidity in the cardiovascular, renal, and hepatic systems, as well as obesity (Lu et al., 2024). Despite the availability of several glucose-lowering medications, many patients struggle to maintain long-term glycemic control, especially when hyperglycemia is combined with obesity and a broader cardiometabolic risk (Vora et al., 2024). In this situation, GLP-1 receptor agonists (GLP-1RAs) have emerged as an important component of modern T2D care due to their ability to lower Hemoglobin A1c (HbA1c), reduce body weight, and provide clinically significant cardiometabolic effects (Raza et al., 2024). American Diabetes Association (ADA) Standards of Care 2026 prioritized GLP-1-based medicines in treatment algorithms, particularly for people with T2D and obesity, cardiovascular disease, or chronic kidney disease.

Despite these benefits, patient response to GLP-1RA medication is substantially varied. Some patients report significant HbA1c reductions and weight loss, while others exhibit just minor improvement or apparent therapeutic failure (Cardoso et al., 2026). This variability has significant clinical implications because GLP-1RAs are increasingly being utilized not only for glycemic control, but also for long-term risk reduction and metabolic disease management. As a result, one of the most pressing unmet needs in care for diabetes is determining which individuals are most likely to respond well and which are more likely to stay poorly controlled despite therapy (Moiz et al., 2025).

The biological cause for this heterogeneity is most likely complex. Baseline glycemic severity, obesity, treatment history, and comorbidity burden all have an impact on therapy results, but these factors may not fully account for the large variations seen in clinical practice (Heni et al., 2025). Pharmacogenomics offers a possible additional rationale. Because GLP-1RAs operate directly on the GLP-1 receptor, hereditary variations in GLP1R may affect receptor signaling, downstream metabolic consequences, and, ultimately, therapeutic response (Dawed et al., 2023). Recent pharmacogenomic investigations and reviews support the hypothesis that genetic variation in GLP1R and related pathways may contribute to interindividual variability in GLP-1RA efficacy (Eghbali et al., 2024). However, this field is still evolving and has not yet been translated into routine clinical decision-making.

At the same time, the response to medication may indicate a broader genetic sensitivity to T2D rather than just target-specific variation (Young et al., 2023). Polygenic risk, which includes hereditary factors such as β-cell malfunction, insulin resistance, and disease severity, may explain why some people respond better than others (Prasad et al., 2025). An integrative paradigm that incorporates longitudinal clinical phenotyping, GLP1R variation, and polygenic risk may thus be more useful than clinical or genetic data alone (Guo et al., 2025).

These concepts provide the foundation for the current investigation. A therapeutically useful assessment of this heterogeneity necessitates comparing HbA1c levels before and after treatment in the same individuals, as well as systematically characterizing anthropometric, metabolic, hepatic, and genetic variables that may influence response. (Umapathysivam et al., 2026). Accordingly, we designed this study as an integrative clinical and pharmacogenomic analysis of GLP-1RA therapy in T2D, with a focus on defining response variability, identifying patient-level factors associated with differential treatment effects, and developing a framework for more precise stratification of people receiving GLP-1RA therapy.

## Methods

### Study design and response stratification

We conducted a retrospective cohort study using electronic health record (EHR)-linked biobank data from version 7 of the All of Us research workbench, with data collected through Oct 2023.

The whole procedure involved identifying patients with T2D who initiated GLP-1RA medications and analyzing available clinical, laboratory, and genetic data, followed by stratifying treatment response using longitudinal HbA1c measurements obtained before and after therapy initiation.

For response analysis, the study population was restricted to individuals with HbA1c data available in both the pre-treatment and post-treatment periods, allowing within-person assessment of glycemic change after GLP-1RA therapy. The treatment index date was established as the beginning of GLP-1RA medication. HbA1c levels measured prior to this date were categorized as pre-treatment, while those measured after this date were classified as post-treatment. Mean HbA1c values for each participant were determined individually before and after therapy to summarize overall glycemic status.

Response stratification was based on the absolute reduction in mean HbA1c, which was computed as the difference between mean pre-treatment and mean post-treatment HbA1c. Patients were defined as good responders if their HbA1c reduction was ≥2.5 percentage point and poor responders if it was <0.5 percentage point (Overgaard et al., 2021; Zhu et al., 2024). The primary comparison analyses focused on good and poor responders to capture the phenotypic extremes of therapeutic benefit and limited response. After response groups were assigned, demographic, anthropometric, metabolic, hepatic, and genetic variables were incorporated into the analytical framework, including body mass index (BMI), blood pressure, fasting glucose, lipid profile, liver function tests, common GLP1R variations, and T2D polygenic risk scores. This strategy allowed for a comprehensive investigation of the clinical and genetic factors that contribute to variability in GLP-1RA response.

### Clinical and biochemical variables

Clinical and laboratory phenotyping was carried out using demographic, anthropometric, hemodynamic, metabolic, hepatic, and genetic variables that were integrated into the study workflow. Demographic characteristics included age, gender, and race/ethnicity. BMI, systolic and diastolic blood pressure were among the clinical and anthropometric measurements taken. HbA1c and fasting glucose were used as glycemic measures; total cholesterol, Low-Density Lipoprotein (LDL) cholesterol, High Density Lipoprotein (HDL) cholesterol, and triglycerides as lipid traits; and alanine aminotransferase, aspartate aminotransferase, alkaline phosphatase, and gamma-glutamyl transferase as liver-related biomarkers.

For treatment-response analyses, HbA1c served as the primary glycemic marker and was used for longitudinal response stratification before and after GLP-1 receptor agonist therapy. Fasting glucose was assessed as an additional indicator of glycemic state. BMI and blood pressure were added to capture the broader clinical and cardiometabolic response, while lipid and liver biomarker panels were used to assess related metabolic and hepatic alterations associated with medication response.

These factors were combined to give a comprehensive phenotypic framework for evaluating GLP-1RA response, allowing comparison of glycemic improvement with concomitant changes in obesity-related, cardiometabolic, hepatic, and hereditary risk.

### Polygenic Risk Score

A T2D polygenic risk score (PRS) was utilized to assess inherited genetic susceptibility to T2D and compare the overall genetic load among GLP-1 receptor agonist responder groups. In individuals with accessible genotype data, the PRS was calculated using weighted risk alleles from established T2D-associated variations discovered in previous genome-wide association studies. After routine genotyping quality control, the PRS was converted to a z-score for subsequent analysis. PRS building was achieved using PRSice (Choi and O’Reilly, 2019) and LDpred2 (Privé et al., 2021), implemented in R.

### Mendelian Randomization Analysis

Mendelian randomization (MR) analysis was performed to evaluate the potential directional relationship between genetically proxied GLP-1RA response and related metabolic traits, including HbA1c, BMI, triglycerides, and liver biomarkers. Common GLP1R-associated variants identified in the pharmacogenomic analysis were used as instrumental variables. SNP-specific effect estimates were harmonized across exposure and outcome datasets, and MR was performed using an inverse-variance weighted framework as the primary method. Sensitivity analyses were conducted using complementary approaches, including MR-Egger and weighted median models, where applicable. MR analyses and visualization were performed using TwoSampleMR (Hermani et al., 2018) and related packages implemented in R. The resulting SNP-level scatter plots were used to compare directional genetic effects across good- and poor-response groups.

### AlphaGenome study

To facilitate functional interpretation of prioritized noncoding variants, regulatory effect prediction was tested using AlphaGenome (Google DeepMind) (Avsec et al., 2026), a sequence-to-function deep learning model that forecasts the effects of DNA variants on multiple regulatory outputs based on long genomic sequence context. AlphaGenome combines long-range sequence information with base-pair-resolution prediction across a variety of regulatory modalities, and it was utilized as an in silico annotation framework to highlight potentially functional variants for downstream analysis.

## Results

### Cohort Overview and Response Classification

The analytical workflow first identified individuals with GLP-1RA exposure and available longitudinal HbA1c measurements. To evaluate treatment response in a clinically meaningful manner, response classification was restricted to individuals with HbA1c data available in both the pre-treatment and post-treatment periods, allowing within-person comparison of glycemic change after therapy initiation. The final response cohort included 3,194 good responders and 2,590 poor responders.

Response was categorized based on the change in HbA1c after GLP-1RA treatment. HbA1c levels obtained prior to treatment commencement were classed as pre-treatment values, whereas those assessed after initiation were classified as post-treatment values. The mean HbA1c was then determined independently for each patient’s pre- and post-treatment periods, and absolute HbA1c decrease was defined as the difference between the mean pre-treatment HbA1c and the mean post-treatment HbA1c. Patients were classified as good responders if their HbA1c reduction was ≥2.5 percentage point and poor responders if it was <0.5 percentage point (**Figure. 1**). Primary comparative analyses focused on the good- and poor-response groups to capture the phenotypic extremes of therapeutic benefit and limited response.

**Figure 1.**
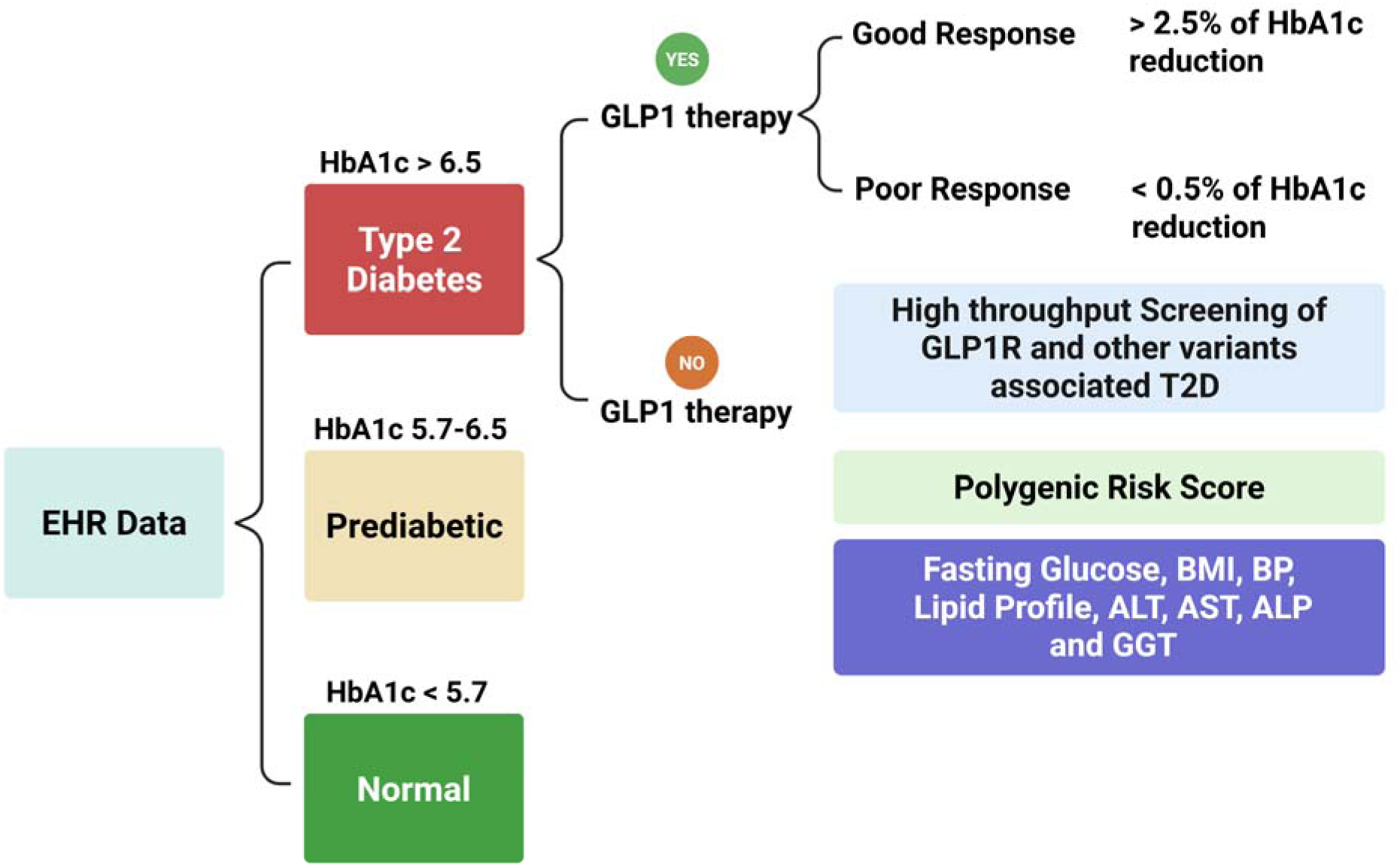
Study workflow for GLP-1RA response analysis in type 2 diabetes. EHR data were stratified by baseline HbA1c into normal, prediabetes, and type 2 diabetes. GLP-1RA-treated patients with type 2 diabetes were classified as good responders (>2.5% HbA1c reduction) or poor responders (<0.5% HbA1c reduction). Downstream analyses included screening of GLP1R and other T2D-associated variants, polygenic risk score analysis, and evaluation of metabolic and clinical traits.

### Demographic Characteristics of Good and Poor Responders

Good responders were younger than poor responders, with median age 55.2 years (range 29–79) versus 58.6 years (31–82), respectively. The proportion of women was similar in the two groups, accounting for 53.5% of good responders and 51.0% of poor responders (**Table 1**).

**Table 1.** Demographic characteristics of good and poor GLP-1RA responders. Values are median (range) or n (%).

| Characteristic | Good Response (N = 3,194) | Poor Response (N = 2,590) |
| --- | --- | --- |
| **Demographics** |  |  |
| **Age, years** | 55.2 (29–79) | 58.6 (31–82) |
| **Female sex** | 1,710 (53.5%) | 1,320 (51.0%) |
| **Race/Ethnicity** |  |  |
| **White** | 1,690 (52.9%) | 1,290 (49.8%) |
| **Black/African American** | 470 (14.7%) | 460 (17.8%) |
| **Asian** | 210 (6.6%) | 150 (5.8%) |
| **Hispanic/Latino** | 640 (20.0%) | 560 (21.6%) |
| **Other/Unknown** | 184 (5.8%) | 130 (5.0%) |

Race and ethnicity distributions were broadly comparable across response groups, although modest differences were observed. Among good responders, 52.9% were White, 14.7% Black/African American, 6.6% Asian, 20.0% Hispanic/Latino, and 5.8% Other/Unknown. Among poor responders, the corresponding proportions were 49.8%, 17.8%, 5.8%, 21.6%, and 5.0%, respectively. Overall, poor responders tended to be slightly older and included a somewhat higher proportion of Black/African American participants, whereas good responders included slightly higher proportions of White and Asian participants (**Table 1**).

These data indicate that the good- and poor-response groups were generally similar demographically, with age representing the most evident difference between groups.

### Glycemic Response to GLP-1RA Therapy

Glycemic response differed markedly between the two groups after GLP-1RA therapy. In good responders, HbA1c decreased from 9.2% before treatment to 6.3% after treatment, corresponding to an absolute HbA1c reduction of 2.9 percentage points. In contrast, poor responders showed only a modest decline in HbA1c, from 8.4% to 8.1%, with an absolute reduction of 0.3 percentage points. The difference in HbA1c reduction between groups was highly significant (P < 0.001) (**Table 2**).

**Table 2.** Clinical, metabolic, and genetic characteristics before and after GLP-1RA therapy in good and poor responders. Values are median (range) or n (%). P values compare differences between response groups.

| **Characteristic** | **Good Response (N=3,194) Before** | **Good Response After** | **Poor Response (N=2,590) Before** | **Poor Response After** | **P value\*** |
| --- | --- | --- | --- | --- | --- |
| **Anthropometric & Clinical** |  |  |  |  |  |
| BMI, kg/m<sup>2</sup> | 34.1 (22.4–51.8) | 32.6 (21.8–49.3) | 36.2 (23.1–55.6) | 35.8 (22.9–54.2) | <0.001 |
| Systolic BP, mmHg | 132 (102–178) | 124 (98–166) | 136 (104–182) | 134 (102–178) | <0.001 |
| Diastolic BP, mmHg | 78 (58–102) | 74 (56–96) | 80 (60–106) | 79 (58–104) | 0.002 |
| **Glycemic Parameters** |  |  |  |  |  |
| HbA1c, % | 9.2 (7.1–13.8) | 6.3 (4.9–8.7) | 8.4 (6.8–12.6) | 8.1 (6.7–11.9) | <0.001 |
| Absolute HbA1c reduction, % | — | 2.9 (2.5–5.6) | — | 0.3 (0.0–0.5) | <0.001 |
| Fasting glucose, mg/dL | 186 (102–412) | 118 (82–212) | 171 (96–398) | 160 (94–356) | <0.001 |
| **Lipid Profile** |  |  |  |  |  |
| Total cholesterol, mg/dL | 182 (104–312) | 168 (98–286) | 190 (110–338) | 186 (106–324) | 0.001 |
| LDL-C, mg/dL | 103 (42–212) | 94 (38–188) | 110 (48–228) | 108 (44–214) | 0.002 |
| HDL-C, mg/dL | 45 (22–88) | 48 (24–92) | 43 (20–82) | 44 (21–84) | 0.008 |
| Triglycerides, mg/dL | 168 (54–612) | 142 (50–480) | 186 (60–745) | 178 (58–710) | <0.001 |
| **Liver Function Tests** |  |  |  |  |  |
| ALT, U/L | 32 (9–148) | 27 (8–120) | 38 (11–186) | 36 (10–174) | <0.001 |
| AST, U/L | 28 (10–122) | 24 (9–98) | 31 (12–140) | 30 (11–132) | 0.001 |
| ALP, U/L | 92 (38–242) | 88 (36–210) | 101 (42–268) | 98 (40–252) | <0.001 |
| GGT, U/L | 44 (12–310) | 36 (10–244) | 58 (15–402) | 54 (14–388) | <0.001 |
| **Genetic /**<br>**Translational**<br>**Variables** |  |  |  |  |  |
| T2D Polygenic Risk<br>Score (z-score) | 0.21 (-2.8–3.1) | — | 0.38 (-2.9–3.3) | — | <0.001 |
| Any GLP1R coding<br>variant carrier | 256 (8.0%) | — | 262 (10.1%) | — | 0.006 |
| Any GLP1R variant<br>(coding +<br>noncoding) | 612 (19.2%) | — | 590 (22.8%) | — | 0.001 |

A similar tendency was seen with fasting glucose. Fasting glucose in good responders fell from 186 mg/dL before therapy to 118 mg/dL after treatment. Poor responders had a moderate reduction in fasting glucose from 171 mg/dL to 160 mg/dL (P < 0.001). These findings suggest a distinct divergence in glycemic trends following GLP-1RA medication. Good responders demonstrated significant reduction in both HbA1c and fasting glucose, but poor responders had consistently elevated glycemic parameters after treatment (**Table 2**).

### Anthropometric and Hemodynamic Changes After Therapy

Anthropometric and blood pressure measures also differed substantially between response groups after GLP-1RA therapy. Among good responders, BMI decreased from 34.1 kg/m² before treatment to 32.6 kg/m² after treatment. In poor responders, the reduction was smaller, from 36.2 kg/m² to 35.8 kg/m² (P < 0.001). A similar pattern was observed for blood pressure. In good responders, systolic blood pressure declined from 132 mmHg before therapy to 124 mmHg after therapy, whereas in poor responders it changed only modestly, from 136 mmHg to 134 mmHg (P < 0.001). Diastolic blood pressure also improved more in good responders, decreasing from 78 mmHg to 74 mmHg, compared with a smaller reduction from 80 mmHg to 79 mmHg in poor responders (P = 0.002) (**Table 2**).

Thus, good responders not only showed better glycemic improvement but also demonstrated greater reductions in body weight–related and hemodynamic measures, whereas poor responders had only limited improvement in these parameters after therapy

### Lipid and Liver Biomarker Changes According to Response Status

Changes in lipid and liver-related biomarkers differed consistently between good and poor responders after GLP-1RA therapy. Good responders showed broader improvement across the lipid profile, with total cholesterol decreasing from 182 to 168 mg/dL, LDL cholesterol from 103 to 94 mg/dL, and triglycerides from 168 to 142 mg/dL. HDL cholesterol increased from 45 to 48 mg/dL. In contrast, poor responders showed only modest lipid changes, with total cholesterol decreasing from 190 to 186 mg/dL, LDL cholesterol from 110 to 108 mg/dL, triglycerides from 186 to 178 mg/dL, and HDL cholesterol increasing only slightly from 43 to 44 mg/dL. Differences between groups were significant for all lipid measures, including total cholesterol (P = 0.001), LDL cholesterol (P = 0.002), HDL cholesterol (P = 0.008), and triglycerides (P < 0.001) (**Table 2**).

A similar pattern was observed for liver-related biomarkers. In good responders, alanine aminotransferase decreased from 32 to 27 U/L, aspartate aminotransferase from 28 to 24 U/L, alkaline phosphatase from 92 to 88 U/L, and gamma-glutamyl transferase from 44 to 36 U/L. By comparison, poor responders showed much smaller changes, with alanine aminotransferase decreasing from 38 to 36 U/L, aspartate aminotransferase from 31 to 30 U/L, alkaline phosphatase from 101 to 98 U/L, and gamma-glutamyl transferase from 58 to 54 U/L. Group differences were significant for alanine aminotransferase (P < 0.001), aspartate aminotransferase (P = 0.001), alkaline phosphatase (P < 0.001), and gamma-glutamyl transferase (P < 0.001) (**Table 2**).

Overall, good responders demonstrated more favorable changes in both lipid and hepatic biomarker profiles, whereas poor responders showed only limited improvement after therapy, consistent with a broader pattern of reduced metabolic benefit in the poor-response group.

### Polygenic risk burden and GLP1R variant enrichment in poor responders

To assess whether inherited susceptibility contributed to treatment heterogeneity, we compared polygenic and locus-specific genetic features between response groups. Poor responders had a higher T2D polygenic risk score than good responders (0.38 vs. 0.21, P < 0.001), suggesting greater global inherited diabetes burden. Poor responders were also more likely to carry any GLP1R coding variant (10.1% vs. 8.0%, P = 0.006) and any GLP1R variant overall, including coding and noncoding variants (22.8% vs. 19.2%, P = 0.001). These findings support the contribution of both broader inherited T2D susceptibility and target-specific pharmacogenomic variation to GLP-1RA response heterogeneity.

### Common GLP1R variants show differential distribution across response groups

Variant-level analyses identified several common GLP1R loci with distinct response-related patterns. The frequency of rs2268650 carriers was substantially higher among poor responders than good responders (20.23% vs. 10.14%). Among good-response carriers, mean HbA1c improved from 8.77 before therapy to 7.26 after therapy, whereas in poor-response carriers HbA1c increased from 8.12 to 8.95. A similar pattern was observed for rs2003132, which was more frequent in poor responders (23.24% vs. 12.90%) and was associated with worsening post-treatment HbA1c in poor responders (8.02 to 9.12) but improvement in good responders (9.02 to 7.82) (**Table 3**).

**Table 3.** Common GLP1R variant carrier frequency and HbA1c values before and after GLP-1 therapy in good and poor responders.

| **Variant** | **Good Response Carriers, n (%)** | **Good HbA1c Before** | **Good HbA1c After** | **Poor Response Carriers, n (%)** | **Poor HbA1c Before** | **Poor HbA1c After** |
| --- | --- | --- | --- | --- | --- | --- |
| rs2268650 | 324 (10.14%) | 8.77 | 7.26 | 524 (20.23%) | 8.12 | 8.95 |
| rs10305439 | 150 (4.70%) | 8.77 | 7.23 | 123 (4.75%) | 8.07 | 8.82 |
| rs5875653 | 221 (6.92%) | 8.92 | 7.54 | 232 (8.96%) | 8.21 | 8.89 |
| rs6906372 | 431 (13.49%) | 8.88 | 7.63 | 182 (7.03%) | 7.91 | 8.90 |
| rs2003132 | 412 (12.90%) | 9.02 | 7.82 | 602 (23.24%) | 8.02 | 9.12 |

Other variants also showed response-specific distributions. rs5875653 was more common in poor responders (8.96% vs. 6.92%) and was associated with poorer post-treatment glycemic control in that group. In contrast, rs6906372 was more common in good responders (13.49% vs. 7.03%), suggesting that some common variants may be associated with more favorable GLP-1RA response. Carrier frequencies for rs10305439 were similar in both groups, although HbA1c improved more substantially among good responders than poor responders (**Table 3**). Collectively, these results indicate that common GLP1R variants differ not only in frequency across response groups but also in the direction and magnitude of associated glycemic trajectories.

### Mendelian Randomization Analysis

MR analyses were utilized to investigate the directional association between genetically proxied GLP-1RA responsiveness and related metabolic variables in both good and poor response groups. The general pattern of the MR scatter plots revealed that SNP-level effects differed between response classes, lending credence to the notion that the genetic architecture of GLP-1RA response goes beyond glycemic change.

The HbA1c MR scatter plots showed a clear directional contrast between response groups. In good responders, all prioritized variants were associated with negative HbA1c effects, consistent with a favorable glycemic profile. In poor responders, the same variants clustered in the positive HbA1c effect range, indicating an unfavorable glycemic pattern. Among individual loci, rs2003132 showed the strongest separation between groups, with the most positive HbA1c effect in poor responders and the negative effect in poor responders. Similar group divergence was observed for rs6906372, rs5875653, rs2268650, and rs10305439 (**Figure 2A**).

**Figure 2.**
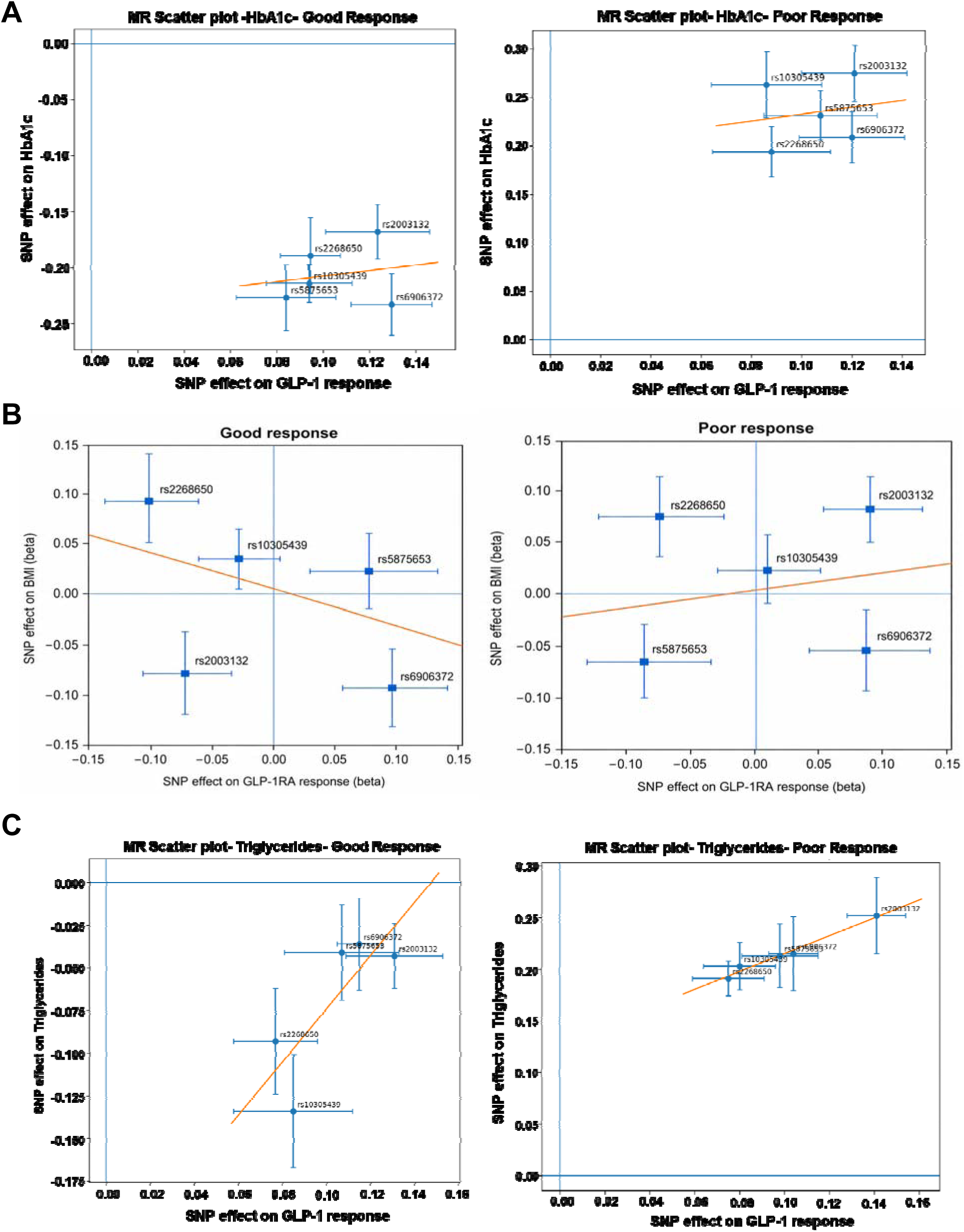
Mendelian randomization scatter plots comparing SNP effects on GLP-1RA response and metabolic traits in good and poor responders. (A) HbA1c, (B) BMI, and (C) triglycerides. Each point represents a prioritized variant, with horizontal and vertical error bars indicating uncertainty in SNP effects on GLP-1RA response and the corresponding metabolic trait, respectively. The orange line indicates the fitted MR trend. Across all three traits, good and poor responders showed distinct directional patterns, supporting differential genetic architecture of GLP-1RA response across glycemic, anthropometric, and lipid phenotypes.

The BMI MR scatter plots also showed a clear contrast between response groups. In good responders, the fitted regression line had a negative slope, suggesting that variants associated with better GLP-1RA response were directionally linked to lower BMI effects. In poor responders, the fitted line showed a positive slope, indicating a less favorable relationship between SNP effects on GLP-1RA response and BMI. At the variant level, rs2268650 and rs10305439 showed positive BMI effects in both groups, whereas rs2003132 and rs6906372 showed negative BMI effects. rs5875653 was positive in good responders but negative in poor responders (**Figure 2B**).

The triglyceride MR scatter plots showed a strong directional contrast between response groups. In good responders, all prioritized variants were associated with negative triglyceride effects, indicating a favorable lipid-related profile. In poor responders, the same variants clustered in the positive triglyceride effect range, consistent with an unfavorable triglyceride pattern.The fitted regression line was positive in both groups, but the overall SNP distribution was clearly shifted downward in good responders and upward in poor responders. Among individual loci, rs2003132 showed the most pronounced positive triglyceride effect in poor responders, whereas rs10305439 and rs2268650 showed the strongest negative triglyceride effects in good responders (**Figure 2C**).

For ALT, the MR scatter plot showed a clear separation between response groups. In good responders, SNP effects were distributed around the null, with some variants showing modest positive effects and others negative effects on ALT. In contrast, in poor responders, all prioritized variants clustered in the positive ALT effect range, indicating a consistent unfavorable hepatic pattern. The fitted regression line was positive in both groups, but the poor-response group showed an overall upward shift in SNP effects compared with good responders (**Figure 3A**).

**Figure 3.**
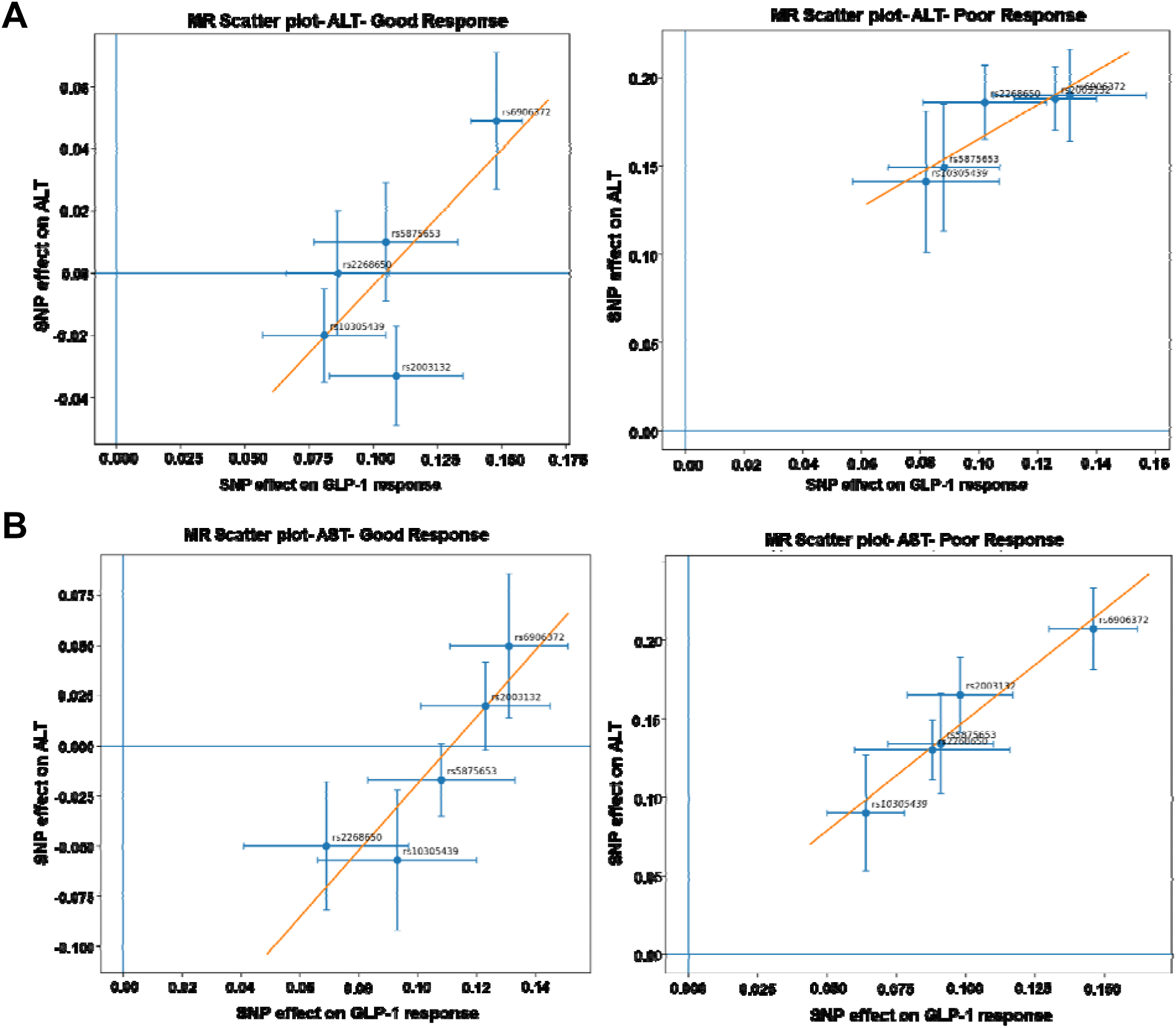
Mendelian randomization scatter plots comparing SNP effects on GLP-1RA response and liver-related biomarkers in good and poor responders. (A) ALT and (B) AST. Each point represents a prioritized variant, with horizontal and vertical error bars indicating uncertainty in SNP effects on GLP-1RA response and the corresponding hepatic trait, respectively. The orange line indicates the fitted MR trend. Good and poor responders showed distinct directional patterns for both ALT and AST, supporting differential genetic associations between GLP-1RA response and liver-related metabolic traits.

For AST, a similar pattern was observed. In good responders, SNP effects were more heterogeneous and largely centered around the null, with several variants showing negative or only modest positive AST effects. In poor responders, all variants were shifted into the positive AST effect range, again indicating an adverse hepatic profile. The positive fitted regression line in the poor-response group was steeper and more clearly separated from the good-response pattern (**Figure 3B**).

At the variant level, rs6906372 showed the strongest positive ALT and AST effects in poor responders, while rs2003132 and rs10305439 also contributed to the upward shift in hepatic biomarker effects. In good responders, the same loci showed weaker, mixed, or even negative effects, particularly for AST.

### AlphaGenome Variant Prediction

To further assess the functional relevance of prioritized GLP1R-associated loci, we performed in silico regulatory interpretation for rs2268650 and rs2003132, which showed significant effects in poor response population. In the AlphaGenome-based pancreas regulatory tracks, both variants showed predicted local effects on chromatin-associated signals at the variant-centered region, supporting a potential regulatory role rather than a purely marker-level association.

Sequence-based prediction of pancreatic ATAC signal for rs2268650 was evaluated across a ±500 bp region centered on the variant. The observed allele-specific difference at the variant site indicates that rs2268650 is predicted to modify local chromatin accessibility, supporting a potential regulatory role rather than a neutral association (**Figure. 4A**). Pancreatic DNase signal prediction showed a pronounced allele-dependent effect centered at rs2268650, indicating that the alternate allele may alter local chromatin accessibility and potentially influence nearby regulatory activity, including transcription factor binding (**Figure. 4B**). Consistent with this, pancreatic eQTL analysis showed a significant genotype-dose association with GLP1R expression, with progressively lower expression from CC to CA to AA genotypes, supporting the A allele as a candidate regulatory allele associated with reduced receptor expression (**Figure. 4C**).

**Figure 4.**
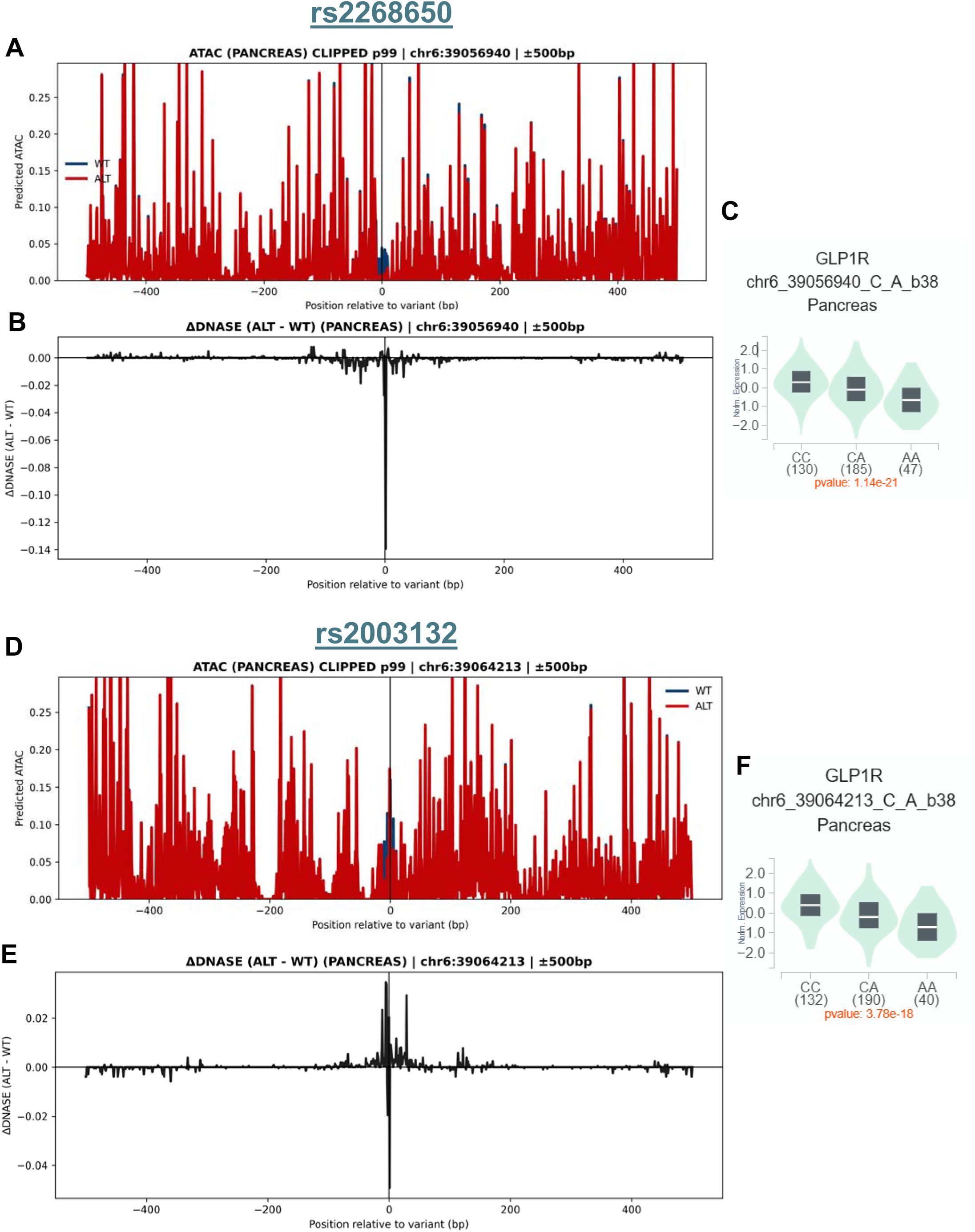
Functional annotation of prioritized GLP1R variants associated with GLP-1RA response. For rs2268650 (**A–C**) and rs2003132 (**D–F**), sequence-based pancreas regulatory predictions are shown for ATAC signal (**A, D**) and allele-specific DNase difference (**B, E**) across a ±500 bp region centered on the variant. Both variants demonstrated allele-dependent effects on local chromatin accessibility and regulatory signal. Pancreatic eQTL analyses (**C, F**) further showed significant genotype-dependent differences in GLP1R expression, with lower expression associated with the alternate allele for both variants, supporting rs2268650 and rs2003132 as candidate functional regulatory loci.

Functional annotation of rs2003132 supported a regulatory role at the GLP1R locus. Sequence-based prediction of pancreatic ATAC signal demonstrated allele-dependent differences across the variant-centered region, consistent with altered local chromatin accessibility (**Figure. 4D**). Pancreatic DNase signal prediction likewise showed a distinct allele-specific effect centered at rs2003132, suggesting that the alternate allele may perturb local regulatory activity (**Figure. 4E**) Consistent with these findings, pancreatic eQTL analysis showed a significant genotype-dependent decrease in GLP1R expression from CC to CA to AA genotypes, indicating that the A allele is associated with lower pancreatic GLP1R expression (**Figure. 4F**) Together, these data support rs2003132 as a candidate functional regulatory variant that may contribute to heterogeneity in GLP-1RA response.

Notably, both rs2268650 and rs2003132 were enriched among poor responders in the clinical-pharmacogenomic analysis and were associated with less favorable post-treatment HbA1c profiles. Taken together, the concordance between the response association, AlphaGenome regulatory prediction, and pancreatic eQTL signal supports the interpretation that these variants may contribute to GLP-1RA response heterogeneity through reduced GLP1R regulatory activity and lower receptor expression in pancreas.

## Discussion

In this integrative clinical and pharmacogenomic analysis of GLP-1RA response in T2D, we observed that treatment heterogeneity was not confined to HbA1c alone, but extended across body weight, blood pressure, lipid traits, liver-related biomarkers, polygenic risk burden, and GLP1R variation. Good responders showed a substantially larger HbA1c reduction than poor responders, together with broader improvement in BMI, fasting glucose, blood pressure, triglycerides, and liver enzymes. Poor responders, by contrast, had persistently adverse metabolic profiles after treatment, higher T2D polygenic risk score, and greater enrichment of GLP1R variants.

This interpretation is clinically relevant because GLP-1–based therapies now occupy a prominent place in contemporary diabetes management, particularly for people with T2D and obesity, cardiovascular disease, or chronic kidney disease (Sreenivasan et al., 2024). Current ADA Standards of Care emphasize GLP-1–based therapy not only for glucose lowering but also for favorable weight and cardiometabolic effects (Rivera et al., 2024). In that context, understanding why some patients derive substantial benefit while others do not has become increasingly important for treatment selection and sequencing.

Our findings are consistent with recent real-world reports showing that response to GLP-1RA therapy is heterogeneous across individuals with T2D (Marassi et al., 2025). A 2025 real-world study of liraglutide, semaglutide, and dulaglutide reported substantial variability in both HbA1c and body-weight response after treatment initiation, supporting the concept that average treatment effects may obscure clinically meaningful subgroups (Heni et al., 2025). The broader metabolic separation observed in our cohort extends that literature by suggesting that heterogeneity in glycemic response may parallel heterogeneity in lipid, blood pressure, and hepatic biomarker response.

An important aspect of our analysis is the observation that poor responders carried a higher T2D polygenic risk burden. Although polygenic risk scores are not yet part of routine therapeutic decision-making, this finding is biologically plausible. Greater inherited burden may reflect more severe underlying defects in glycemic regulation, insulin resistance, or β-cell reserve, which could limit the degree of benefit achievable with incretin-based therapy (Lee et al., 2024). More broadly, this result supports the growing view that precision diabetes treatment will likely require integration of baseline phenotype with inherited susceptibility rather than reliance on clinical variables alone.

Our locus-specific results also support a role for receptor-level pharmacogenomics. Prior pharmacogenomic studies have suggested that genetic variability at GLP1R may influence glycemic response to GLP-1RAs, although available reports remain relatively limited and effect estimates have varied across cohorts. In this context, the enrichment of GLP1R variants among poor responders in our study is directionally consistent with the emerging literature and strengthens the rationale for evaluating receptor-region variation as one determinant of treatment-response heterogeneity.

Among the variants examined, rs2268650 and rs2003132 were the most consistently prioritized. These loci were enriched in poor responders and were further supported by regulatory annotation and pancreatic eQTL evidence. Although these data do not establish causality, the convergence of clinical association, in silico regulatory prediction, and genotype-dependent pancreatic GLP1R expression supports the possibility that altered regulation at the receptor locus contributes to reduced therapeutic response in a subset of patients. In this regard, our data provides a basis for future functional studies rather than definitive mechanistic proof.

The Mendelian randomization analyses should likewise be interpreted cautiously. Rather than establishing direct causality, these analyses support the possibility that the genetic architecture associated with GLP-1RA response may relate to multiple downstream traits, including glycemic, lipid, hepatic, and anthropometric phenotypes. This broader pattern is compatible with the known pleiotropic effects of GLP-1RA therapy and suggests that poor response may represent a wider metabolic phenotype rather than isolated glycemic nonresponse. This study has limitations. First, the analysis was observational, and the response groups were defined within a clinical rather than randomized framework. Second, while our functional annotation strengthens prioritization of selected loci, experimental validation remains necessary. Third, pharmacogenomic studies of GLP-1RA response are still evolving, and replication across independent and ancestrally diverse cohorts will be important before any translational application. Nonetheless, the consistency between our clinical, pharmacogenomic, and regulatory findings supports the relevance of this integrated framework for future work.

Overall, our results support the idea that response to GLP-1RA therapy in T2D is influenced by both baseline clinical-metabolic state and inherited biology. From a translational perspective, the major implication is that clinically useful response stratification may ultimately require combining routine clinical phenotyping with pharmacogenomic information, particularly as GLP-1–based therapies continue to expand in diabetes care.

## Conclusion

In conclusion, GLP-1RA response heterogeneity in T2D appears to reflect more than differences in HbA1c change alone. In our cohort, poor response was accompanied by a less favorable metabolic profile, higher polygenic risk burden, and enrichment of selected GLP1R variants, with additional regulatory support for rs2268650 and rs2003132. These findings support further evaluation of integrated clinical and pharmacogenomic approaches to improve understanding of GLP-1RA response variability and to inform future precision-treatment strategies in type 2 diabetes.

## Acknowledgments

We gratefully acknowledge All of Us participants for their contributions, without whom this research would not have been possible. We also thank the National Institutes of Health’s All of Us Research Program for making available the participant data examined in this study. The authors thank NYU Langone Hospital—Long Island for providing computational resources. AI-based tools were used solely to improve English language and grammar.

## Data Availability

This study used data from the National Institutes of Health All of Us Research Program’s Controlled Tier Dataset, version 7, available to authorized users through the Researcher Workbench. Participant-level data cannot be publicly shared by the authors.

## References

Lu X, Xie Q, Pan X, Zhang R, Zhang X, Peng G, et al. Type 2 diabetes mellitus in adults: pathogenesis, prevention and therapy. Signal Transduct Target Ther. 2024;9(1):262. doi:10.1038/s41392-024-01951-9

Vora J, Cherney D, Kosiborod MN, Spaak J, Kanumilli N, Khunti K, et al; CaReMe Global Alliance. Inter-relationships between cardiovascular, renal and metabolic diseases: underlying evidence and implications for integrated interdisciplinary care and management. Diabetes Obes Metab. 2024;26(5):1567–1581. doi:10.1111/dom.15485

Raza FA, Altaf R, Bashir T, Asghar F, Altaf R, Tousif S, et al. Effect of GLP-1 receptor agonists on weight and cardiovascular outcomes: a review. Medicine (Baltimore). 2024;103(44):e40364. doi:10.1097/MD.0000000000040364

American Diabetes Association Professional Practice Committee. 9. Pharmacologic approaches to glycemic treatment: Standards of Care in Diabetes—2026. Diabetes Care. 2026;49(Suppl 1):S183-S215. doi:10.2337/dc26-S009

Cardoso P, Dennis JM, Pearson ER. GLP-1RA precision medicine in people with type 2 diabetes: current insights and future prospects. J Clin Invest. 2026;136(2):e194742. doi:10.1172/JCI194742

Moiz A, Filion KB, Tsoukas MA, Yu OHY, Peters TM, Eisenberg MJ. The expanding role of GLP-1 receptor agonists: a narrative review of current evidence and future directions. EClinicalMedicine. 2025;86:103363. doi:10.1016/j.eclinm.2025.103363

Heni M, Frühwald L, Karges W, Naudorf M, Niemöller K, Pagnia F, et al. Heterogeneity in response to GLP-1 receptor agonists in type 2 diabetes in real-world clinical practice: insights from the DPV register - an IMI-SOPHIA study. Diabetologia. 2025;68(8):1666–1673. doi:10.1007/s00125-025-06448-w

Dawed AY, Mari A, Brown A, McDonald TJ, Li L, Wang S, et al; DIRECT Consortium. Pharmacogenomics of GLP-1 receptor agonists: a genome-wide analysis of observational data and large randomised controlled trials. Lancet Diabetes Endocrinol. 2023;11(1):33-41. doi:10.1016/S2213-8587(22)00340-0

Eghbali M, Alaei-Shahmiri F, Hashemi-Madani N, Emami Z, Mostafavi L, Malek M, et al. Glucagon-like peptide 1 receptor variants and glycemic response to liraglutide: a pharmacogenetics study in Iranian people with type 2 diabetes mellitus. Adv Ther. 2024;41(2):826–836. doi:10.1007/s12325-023-02761-1

Young KG, McInnes EH, Massey RJ, Kahkoska AR, Pilla SJ, Raghaven S, et al; ADA/EASD Precision Medicine in Diabetes Initiative Consortium. Precision medicine in type 2 diabetes: a systematic review of treatment effect heterogeneity for GLP-1 receptor agonists and SGLT2 inhibitors. medRxiv. 2023. doi:10.1101/2023.04.21.23288868

Prasad RB, Hakaste L, Tuomi T. Clinical use of polygenic scores in type 2 diabetes: challenges and possibilities. Diabetologia. 2025;68(7):1361–1374. doi:10.1007/s00125-025-06419-1

Guo B, Cai Y, Kim D, et al. Polygenic risk score for type 2 diabetes shows context-dependent effects across populations. Nat Commun. 2025;16(1):8632. doi:10.1038/s41467-025-63546-4

Umapathysivam MM, Araldi E, Hastoy B, et al. Type 2 diabetes risk alleles in peptidyl-glycine alpha-amidating monooxygenase influence GLP-1 levels and response to GLP-1 receptor agonists. Genome Med. 2026;18(1):40. doi:10.1186/s13073-026-01630-0

All of Us Research Program Investigators, Denny JC, Rutter JL, et al. The "All of Us" Research Program. N Engl J Med. 2019;381(7):668-676. doi:10.1056/NEJMsr1809937

Overgaard RV, Hertz CL, Ingwersen SH, Navarria A, Drucker DJ. Levels of circulating semaglutide determine reductions in HbA1c and body weight in people with type 2 diabetes. Cell Rep Med. 2021;2(9):100387. doi:10.1016/j.xcrm.2021.100387

Zhu X, Fowler MJ, Wells QS, Stafford JM, Gannon M. Predicting responsiveness to GLP-1 pathway drugs using real-world data. BMC Endocr Disord. 2024;24(1):269. doi:10.1186/s12902-024-01798-9

Choi SW, O’Reilly PF. PRSice-2: polygenic risk score software for biobank-scale data. Gigascience. 2019;8(7):giz082. doi:10.1093/gigascience/giz082

Privé F, Arbel J, Vilhjálmsson BJ. LDpred2: better, faster, stronger. Bioinformatics. 2021;36(22-23):5424–5431. doi:10.1093/bioinformatics/btaa1029

Hemani G, Zheng J, Elsworth B, Wade KH, Haberland V, Baird D, et al. The MR-Base platform supports systematic causal inference across the human phenome. eLife. 2018;7:e34408. doi:10.7554/eLife.34408

Avsec Ž, Latysheva N, Cheng J, et al. Advancing regulatory variant effect prediction with AlphaGenome. Nature. 2026;649(8099):1206–1218. doi:10.1038/s41586-025-10014-0

Sreenivasan C, Parikh A, Francis AJ, et al. Evaluating cardiovascular benefits of glucagon-like peptide-1 receptor agonists in type 2 diabetes mellitus: a systematic review. Cureus. 2024;16(8):e66697. doi:10.7759/cureus.66697

Rivera FB, Cruz LLA, Magalong JV, et al. Cardiovascular and renal outcomes of glucagon-like peptide 1 receptor agonists among patients with and without type 2 diabetes mellitus: a meta-analysis of randomized placebo-controlled trials. Am J Prev Cardiol. 2024;18:100679. doi:10.1016/j.ajpc.2024.100679

Marassi M, Cignarella A, Russo GT, et al. Sex differences in the weight response to GLP-1RA in people with type 2 diabetes. A long-term longitudinal real-world study. Pharmacol Res. 2025;219:107866. doi:10.1016/j.phrs.2025.107866

Lee H, Choi J, Kim JI, et al. Higher genetic risk for type 2 diabetes is associated with a faster decline of β-cell function in an East Asian population. Diabetes Care. 2024;47(8):1386–1394. doi:10.2337/dc24-0058

